## Supplementary tables and figures for "ADHD and Autism Spectrum Disorder (ASD) in Childhood: establishing the feasibility and validity of a nation-wide e-cohort"

Table s1: Details on the datasets utilised in this study:

| Dataset | Description | Coverage | Timescale |
| --- | --- | --- | --- |
| The Welsh Demographics Service (WDS) | Register of all individuals who have ever had contact with the NHS or registered with a Welsh GP. | All individuals in Wales | Whole of the study period |
| Welsh Index of Multiple Deprivation (WIMD) | Dataset assigning a deprivation score derived from eight domains including employment, income and education to all Lower Super Output Areas (LSOAs; geographical areas comprising of around 1500 individuals). Individuals are assigned a deprivation index based on the LSOA of their current address in WDS. | All individuals in Wales | Whole of the study period |
| General Practice Database (GPD) | Attendance and clinical information for all primary care contacts. Includes diagnoses, symptoms and prescriptions. | 79% of individuals in Wales from 333/432 GP practices | Whole of the study period |
| Patient Episode Database for Wales (PEDW) | Attendance and clinical information for all NHS Wales hospital admissions (both inpatient and day cases). Includes diagnoses, and specialty. | All individuals in Wales | Whole of the study period |
| Emergency Department Dataset | Administrative and clinical information for all NHS Wales Accident and Emergency Department attendances. | All individuals in Wales | Data available since 2009 |

Full details of these datasets are available at [www.saildatabank.com](http://www.saildatabank.com)

Table s2: List of read codes, ICD-10 codes and prescription codes utilised to identify ADHD and ASD:

| Concept | Read codes and prescription codes | ICD-10 codes |
| --- | --- | --- |
| ADHD | **Eu900, 9Ngp.., Eu9y7, E2E01, EU900, 6A61, 8BPT.., EU901, EU90.., E2E.., Zs91.00, zS91.11, ZS91.12, dc1.., dw1.., dw2.., dw3..,** | [**F90**](http://www.icd10data.com/ICD10CM/Codes/F01-F99/F90-F98/F90-/F90)**, F90.0, F90.1, F90.2, F90.8, F90.9** |
| ASD | **E140, Eu840, Eu841, Eu845, Eu84z-1, 1J9** | **F84.0, F84.5, F84.9** |

Table S3: Cox’s regression analysis: **ADHD associations with outcomes**

|  | **Model 1** | | **Model 2** | | **Model 3** | |
| --- | --- | --- | --- | --- | --- | --- |
|  | **HR** | **95% CI** | **HR** | **95% CI** | **HR** | **95% CI** |
| **Anxiety/depression** |  |  |  |  |  |  |
| **ADHD** | **2.36** | **2.20, 2.53** | **2.36** | **2.20, 2.53** | **2.32** | **2.17, 2.50** |
| Sex |  |  | 0.44 | 0.41, 0.47 | 0.44 | 0.41, 0.47 |
| Deprivation** |  |  |  |  | 1.05 | 1.02, 1.07 |
| **Self-harm** |  |  |  |  |  |  |
| **ADHD** | **5.70** | **5.07, 6.40** | **5.70** | **5.06, 6.39** | **5.52** | **4.91, 6.20** |
| Sex |  |  | 0.53 | 0.47, 0.60 | 0.43 | 0.47, 0.60 |
| Deprivation** |  |  |  |  | 1.12 | 1.08, 1.17 |
| **Alcohol use** |  |  |  |  |  |  |
| **ADHD** | **3.95** | **3.42, 4.56** | **3.95** | **3.42, 4.56** | **3.85** | **3.33, 4.44** |
| Sex |  |  | 1.07 | 0.88, 1.28 | 1.06 | 0.88, 1.28 |
| Deprivation** |  |  |  |  | 1.10 | 1.05, 1.16 |
| **Drug use** |  |  |  |  |  |  |
| **ADHD** | **5.88** | **5.08, 6.80** | **5.88** | **5.09, 6.81** | **5.68** | **4.90, 6.57** |
| Sex |  |  | 1.49 | 1.21, 1.83 | 1.49 | 1.21, 1.83 |
| Deprivation** |  |  |  |  | 1.15 | 1.09, 1.21 |
| Emergency Department**room use** |  |  |  |  |  |  |
| **ADHD** | **1.36** | **1.31, 1.41** | **1.36** | **1.31, 1.41** | **1.34** | **1.30, 1.39** |
| Sex |  |  | 0.96 | 0.92, 1.00 | 0.96 | 0.92, 1.00 |
| Deprivation** |  |  |  |  | 1.04 | 1.02, 1.05 |
| **Any primary care use** |  |  |  |  |  |  |
| **ADHD** | 2.63 | 2.46, 2.80 | 2.62 | 2.46, 2.80 | 2.58 | 2.42, 2.76 |
| Sex |  |  | 0.50 | 0.46, 0.53 | 0.50 | 0.46, 0.53 |
| Deprivation** |  |  |  |  | 1.06 | 1.03, 1.08 |
| **Any hospital use (inc ED)** |  |  |  |  |  |  |
| **ADHD** | 1.36 | 1.31, 1.41 | 1.36 | 1.31, 1.41 | 1.35 | 1.30, 1.40 |
| Sex |  |  | 0.95 | 0.91, 0.99 | 0.95 | 0.91, 0.99 |
| Deprivation** |  |  |  |  | 1.03 | 1.02, 1.05 |

Models incrementally adjusting for covariates. * Age at end of follow up period as time variable; **WIMD quintile

Table s4: Binomial analysis of number of recorded incidents: ADHD and ASD associations with number of Anxiety/depression, self-harm and Emergency Department events

|  | **n** | **B** | **St. error** | **95% CI** |
| --- | --- | --- | --- | --- |
| **ADHD** |  |  |  |  |
| **Number of self-harm events** | 1216 | 0.43 | 0.004 | 0.35, 0.51 |
| **Number of Emergency Department visits** | 14916 | 0.53 | 0.009 | 0.51, 0.55 |
| **ASD** |  |  |  |  |
| **Number of self-harm events** | 484 | 0.53 | 0.06 | 0.42, 0.64 |
| **Number of Emergency Department visits** | 7514 | 0.26 | 0.01 | 0.23, 0.29 |

* Controlling for sex, age at end of follow up, proportion of follow up and deprivation

Table s5: Cox’s regression analysis: **ADHD associations with self-harm using GP or hospital records only**

|  | **Model 1** | | **Model 2** | | **Model 3** | |
| --- | --- | --- | --- | --- | --- | --- |
|  | **HR** | **95% CI** | **HR** | **95% CI** | **HR** | **95% CI** |
| **GP records** |  |  |  |  |  |  |
| **ADHD** | **6.61** | **5.65, 7.72** | **5.72** | **4.88, 6.70** | **5.52** | **4.71, 6.47** |
| Sex |  |  | 0.43 | 0.37, 0.50 | 0.43 | 0.36, 0.50 |
| Deprivation** |  |  |  |  | 1.14 | 1.08, 1.20 |
| **Hospital records** |  |  |  |  |  |  |
| **ADHD** | **7.07** | **5.91, 8.48** | **5.94** | **4.95, 7.13** | **5.76** | **4.80, 6.92** |
| Sex |  |  | 0.44 | 0.37, 0.52 | 0.44 | 0.37, 0.52 |
| Deprivation** |  |  |  |  | 1.12 | 1.05, 1.19 |

Models incrementally adjusting for covariates. * Age at end of follow up period as time variable; ** WIMD quintile

Table s6: Cox’s regression analysis: **ADHD associations with outcomes MALES**

|  | **Model 1** | | **Model 2** | |
| --- | --- | --- | --- | --- |
|  | **HR** | **95% CI** | **HR** | **95% CI** |
| **Anxiety/depression** |  |  |  |  |
| **ADHD** | **2.36** | **2.16, 2.57** | **2.32** | **2.13, 2.54** |
| Deprivation** |  |  | 1.06 | 1.03, 1.09 |
| **Self-harm** |  |  |  |  |
| **ADHD** | **5.75** | **5.00, 6.61** | **5.53** | **4.80, 6.36** |
| Deprivation** |  |  | 1.16 | 1.10, 1.22 |
| **Alcohol use** |  |  |  |  |
| **ADHD** | **3.70** | **3.16, 4.33** | **3.58** | **3.06, 4.19** |
| Deprivation** |  |  | 1.13 | 1.07, 1.20 |
| **Drug use** |  |  |  |  |
| **ADHD** | **5.67** | **4.85, 6.62** | **5.47** | **4.67, 6.39** |
| Deprivation** |  |  | 1.14 | 1.08, 1.21 |
| Emergency Department **use** |  |  |  |  |
| **ADHD** | **1.32** | **1.27, 1.38** | **1.31** | **1.26, 1.36** |
| Deprivation** |  |  | 1.03 | 1.02, 1.04 |
| **Any primary care use** |  |  |  |  |
| **ADHD** | **2.70** | **2.50, 2.92** | **2.65** | **2.45, 2.87** |
| Deprivation** |  |  | 1.07 | 1.04, 1.10 |
| **Any hospital use (inc ED)** |  |  |  |  |
| **ADHD** | **1.32** | **1.27, 1.38** | **1.31** | **1.26, 1.37** |
| Deprivation** |  |  | 1.03 | 1.02, 1.04 |

Models incrementally adjusting for covariates. * Age at end of follow up period as time variable; ** WIMD quintile

Table s7: Cox’s regression analysis: **ADHD associations with outcomes FEMALES**

|  | **Model 1** | | **Model 2** | |
| --- | --- | --- | --- | --- |
|  | **HR** | **95% CI** | **HR** | **95% CI** |
| **Anxiety/depression** |  |  |  |  |
| **ADHD** | **2.35** | **2.09, 2.65** | **2.34** | **2.07, 2.64** |
| Deprivation** |  |  | 1.03 | 0.99, 1.07 |
| **Self-harm** |  |  |  |  |
| **ADHD** | **5.57** | **4.51, 6.88** | **5.50** | **4.45, 6.80** |
| Deprivation** |  |  | 1.05 | 0.98, 1.130 |
| **Alcohol use** |  |  |  |  |
| **ADHD** | **5.45** | **3.84, 7.75** | **5.49** | **3.86, 7.80** |
| Deprivation** |  |  |  |  |
| **Drug use** |  |  |  |  |
| **ADHD** | **7.69** | **5.05, 11.71** | **7.41** | **4.86, 11.30** |
| Deprivation** |  |  | 1.17 | 1.01, 1.35 |
| Emergency Department **use** |  |  |  |  |
| **ADHD** | **1.51** | **1.39, 1.63** | **1.49** | **1.38, 1.61** |
| Deprivation** |  |  | 1.05 | 1.03, 1.08 |
| **Any primary care use** |  |  |  |  |
| **ADHD** | **2.47** | **2.20, 2.77** | **2.44** | **2.18, 2.75** |
| Deprivation** |  |  | 1.04 | 0.99, 1.08 |
| **Any hospital use (inc ED)** |  |  |  |  |
| **ADHD** | **1.52** | **1.41, 1.64** | **1.50** | **1.39, 1.62** |
| Deprivation** |  |  | 1.05 | 1.03, 1.08 |

Models incrementally adjusting for covariates. * Age at end of follow up period as time variable; ** WIMD quintile

Table s8: Cox’s regression – associations between ADHD and outcomes, stratified by Welsh index of multiple deprivation (WIMD):

|  | **Model 1*** |  | **Model 2**** |  |
| --- | --- | --- | --- | --- |
|  | **HR** | **95% CI** | **HR** | **95% CI** |
| **Anxiety/depression** |  |  |  |  |
| WIMD 1: | 2.34 | 1.93, 2.84 | 2.38 | 1.97, 2.89 |
| WIMD 2: | 2.97 | 2.47, 3.57 | 2.92 | 2.43, 3.51 |
| WIMD 3: | 2.41 | 2.04, 2.86 | 2.35 | 1.99, 2.78 |
| WIMD 4: | 2.10 | 1.81, 2.45 | 2.12 | 1.82, 2.47 |
| WIMD 5: | 2.20 | 1.95, 2.94 | 2.21 | 1.96, 2.50 |
| **Self-harm** |  |  |  |  |
| WIMD 1: | 5.17 | 3.61, 7.39 | 5.26 | 3.68, 7.53 |
| WIMD 2: | 10.51 | 7.43, 14.86 | 10.43 | 7.37, 14.75 |
| WIMD 3: | 6.58 | 4.97, 8.71 | 6.41 | 4.85, 8.49 |
| WIMD 4: | 4.72 | 3.72, 5.99 | 4.75 | 3.75, 6.03 |
| WIMD 5: | 4.46 | 3.66, 5.43 | 4.47 | 3.67, 5.44 |
| **Alcohol use** |  |  |  |  |
| WIMD 1: | 5.74 | 3.58, 9.23 | 5.78 | 3.60, 9.28 |
| WIMD 2: | 4.16 | 2.80, 6.14 | 4.15 | 2.81, 6.13 |
| WIMD 3: | 3.19 | 2.37, 4.29 | 3.19 | 2.37, 4.29 |
| WIMD 4: | 4.38 | 3.21, 5.96 | 4.37 | 3.21, 5.96 |
| WIMD 5: | 3.48 | 2.70, 4.49 | 3.48 | 2.70, 4.48 |
| **Drug use:** |  |  |  |  |
| WIMD 1: | 5.40 | 3.50, 8.33 | 5.36 | 3.47, 8.27 |
| WIMD 2: | 8.06 | 5.23, 12.42 | 8.12 | 5.27, 12.51 |
| WIMD 3: | 5.62 | 4.01, 7.88 | 5.65 | 4.03, 7.92 |
| WIMD 4: | 6.61 | 4.74, 9.20 | 6.57 | 4.72, 9.16 |
| WIMD 5: | 4.75 | 3.75, 6.01 | 4.74 | 3.75, 6.01 |
| **ED Use:** |  |  |  |  |
| WIMD 1: | 1.36 | 1.23, 1.50 | 1.36 | 1.23, 1.50 |
| WIMD 2: | 1.35 | 1.23, 1.48 | 1.35 | 1.23, 1.48 |
| WIMD 3: | 1.37 | 1.26, 1.48 | 1.36 | 1.26, 1.48 |
| WIMD 4: | 1.27 | 1.18, 1.37 | 1.27 | 1.18, 1.37 |
| WIMD 5: | 1.38 | 1.29, 1.47 | 1.38 | 1.29, 1.47 |
| **Any primary care use:** |  |  |  |  |
| WIMD 1: | 2.41 | 2.00, 2.89 | 2.44 | 2.03, 2.93 |
| WIMD 2: | 3.29 | 2.77, 3.91 | 3.24 | 2.73, 3.85 |
| WIMD 3: | 2.74 | 2.36, 3.19 | 2.31 | 2.31, 3.12 |
| WIMD 4: | 2.42 | 2.11, 2.79 | 2.44 | 2.12, 2.81 |
| WIMD 5: | 2.42 | 2.16, 2.71 | 2.42 | 2.16, 2.72 |
| **Any hospital use (inc ED)** |  |  |  |  |
| WIMD 1: | 1.36 | 1.24, 1.50 | 1.36 | 1.24, 1.50 |
| WIMD 2: | 1.36 | 1.23, 1.49 | 1.36 | 1.23, 1.49 |
| WIMD 3: | 1.37 | 1.26, 1.49 | 1.37 | 1.26, 1.48 |
| WIMD 4: | 1.28 | 1.18, 1.38 | 1.28 | 1.19, 1.38 |
| WIMD 5: | 1.38 | 1.29, 1.46 | 1.38 | 1.29, 0.99 |

* Unadjusted association with ADHD; ** Association with ADHD, controlling for Sex.

Table s9: Cox’s regression analysis: ADHD associations with outcomes, sample with complete data coverage only

|  | **Model 1** | | **Model 2** | | **Model 3** | |
| --- | --- | --- | --- | --- | --- | --- |
|  | **HR** | **95% CI** | **HR** | **95% CI** | **HR** | **95% CI** |
| **Anxiety/depression** |  |  |  |  |  |  |
| **ADHD** | **2.23** | **2.07, 2.40** | **2.23** | **2.07, 2.40** | **2.20** | **2.04, 2.37** |
| Sex |  |  | 0.43 | 0.39, 0.46 | 0.43 | 0.39, 0.46 |
| Deprivation** |  |  |  |  | 1.05 | 1.02, 1.07 |
| **Self-harm** |  |  |  |  |  |  |
| **ADHD** | **5.46** | **4.83, 6.18** | **5.46** | **4.83, 6.17** | **5.30** | **4.68, 5.99** |
| Sex |  |  | 0.53 | 0.46, 0.60 | 0.53 | 0.46, 0.60 |
| Deprivation** |  |  |  |  | 1.12 | 1.07, 1.17 |
| **Alcohol use** |  |  |  |  |  |  |
| **ADHD** | **3.81** | **3.29, 4.43** | **3.82** | **3.29, 4.43** | **3.71** | **3.19, 4.31** |
| Sex |  |  | 1.06 | 0.87, 1.30 | 1.06 | 0.87, 1.30 |
| Deprivation** |  |  |  |  | 1.11 | 1.05, 1.17 |
| **Drug use** |  |  |  |  |  |  |
| **ADHD** | **5.59** | **4.80, 6.51** | **5.59** | **4.80, 6.51** | **5.39** | **4.62, 6.28** |
| Sex |  |  | 1.52 | 1.22, 1.90 | 1.52 | 1.22, 1.90 |
| Deprivation** |  |  |  |  | 1.14 | 1.08, 1.21 |
| Emergency Department **use** |  |  |  |  |  |  |
| **ADHD** | **1.36** | **1.31, 1.41** | **1.36** | **1.31, 1.41** | **1.35** | **1.30, 1.40** |
| Sex |  |  | 0.94 | 0.91, 0.99 | 0.95 | 0.91, 1.04 |
| Deprivation** |  |  |  |  | 1.03 | 1.02, 1.04 |
| **Any primary care use** |  |  |  |  |  |  |
| **ADHD** | 2.46 | 2.30, 2.63 | 2.46 | 2.30, 2.63 | 2.42 | 2.26, 2.59 |
| Sex |  |  | 0.48 | 0.45, 0.52 | 0.48 | 0.45, 0.52 |
| Deprivation** |  |  |  |  | 1.06 | 1.03, 1.08 |
| **Any hospital use (inc ED)** |  |  |  |  |  |  |
| **ADHD** | 1.36 | 1.32, 1.42 | 1.36 | 1.32, 1.42 | 1.35 | 1.30, 1.40 |
| Sex |  |  | 0.94 | 0.90, 0.98 | 0.94 | 0.90, 0.98 |
| Deprivation** |  |  |  |  | 1.03 | 1.02, 1.04 |

Models incrementally adjusting for covariates. * Age at end of follow up period as time variable; **WIMD quintile

Table S10: Cox’s regression analysis: **ASD associations with outcomes**

|  | **Model 1** | | **Model 2** | | **Model 3** | |
| --- | --- | --- | --- | --- | --- | --- |
|  | **HR** | **95% CI** | **HR** | **95% CI** | **HR** | **95% CI** |
| **Anxiety/depression** |  |  |  |  |  |  |
| **ASD** | **2.11** | **1.91, 2.34** | **2.14** | **1.93, 2.36** | **2.14** | **1.93, 2.63** |
| Sex |  |  | 0.47 | 0.42, 0.52 | 0.47 | 0.42, 0.52 |
| Deprivation** |  |  |  |  | 1.06 | 1.03, 1.10 |
| **Self-harm** |  |  |  |  |  |  |
| **ASD** | **2.93** | **2.45, 3.50** | **2.96** | **2.48, 3.54** | **2.96** | **2.48, 3.54** |
| Sex |  |  | 0.49 | 0.41, 0.59 | 0.50 | 0.41, 0.60 |
| Deprivation** |  |  |  |  | 1.10 | 1.03, 1.17 |
| **Alcohol use** |  |  |  |  |  |  |
| **ASD** | **1.19** | **0.91, 1.55** | **1.19** | **0.91, 1.55** | **1.19** | **0.91, 1.55** |
| Sex |  |  | 1.19 | 0.88, 1.61 | 1.21 | 0.89, 1.63 |
| Deprivation** |  |  |  |  | 1.10 | 1.01, 1.20 |
| **Drug use** |  |  |  |  |  |  |
| **ASD** | **2.21** | **1.66, 2.95** | **2.20** | **1.65, 2.93** | **2.20** | **1.65, 2.94** |
| Sex |  |  | 2.37 | 1.51, 3.73 | 2.42 | 1.54, 3.81 |
| Deprivation** |  |  |  |  | 1.19 | 1.07, 1.31 |
| Emergency Department **use** |  |  |  |  |  |  |
| **ASD** | **0.95** | **0.90, 1.00** | **0.95** | **0.90, 1.00** | **0.95** | **0.90, 1.00** |
| Sex |  |  | 0.99 | 0.93, 1.04 | 0.99 | 0.94, 1.05 |
| Deprivation** |  |  |  |  | 1.05 | 1.04, 1.07 |
| **Any primary care use** |  |  |  |  |  |  |
| **ASD** | 1.99 | 1.81, 2.19 | 2.01 | 1.82, 2.21 | 2.01 | 1.82, 2.22 |
| Sex |  |  | 0.50 | 0.46, 0.56 | 0.51 | 0.46, 0.56 |
| Deprivation** |  |  |  |  | 1.06 | 1.02, 1.10 |
| **Any hospital use (inc ED)** |  |  |  |  |  |  |
| **ASD** | 0.96 | 0.91, 1.01 | 0.96 | 0.91, 1.01 | 0.96 | 0.91, 1.01 |
| Sex |  |  | 0.98 | 0.93, 1.04 | 0.99 | 0.94, 1.04 |
| Deprivation** |  |  |  |  | 1.05 | 1.04, 1.07 |

Models incrementally adjusting for covariates. * Age at end of follow up period as time variable; **WIMD quintile

Table s11: Cox’s regression analysis: **ASD associations with self-harm using GP or hospital records only**

|  | **Model 1** | | **Model 2** | | **Model 3** | |
| --- | --- | --- | --- | --- | --- | --- |
|  | **HR** | **95% CI** | **HR** | **95% CI** | **HR** | **95% CI** |
| **GP records** |  |  |  |  |  |  |
| **ASD** | **3.60** | **2.86, 4.53** | **3.25** | **2.58, 4.10** | **3.26** | **2.58, 4.10** |
| Sex |  |  | 0.40 | 0.31, 0.50 | 0.40 | 0.32, 0.50 |
| Deprivation** |  |  |  |  | 1.09 | 1.002, 1.18 |
| **Hospital records** |  |  |  |  |  |  |
| **ASD** | **3.75** | **2.85, 4.93** | **3.32** | **2.52, 4.37** | **3.32** | **2.52, 4.37** |
| Sex |  |  | 0.40 | 0.30, 0.52 | 0.40 | 0.30, 0.52 |
| Deprivation** |  |  |  |  | 1.03 | 0.93, 1.13 |

Models incrementally adjusting for covariates. * Age at end of follow up period as time variable; ** WIMD quintile

Table s12: Cox’s regression analysis: **ASD associations with outcomes MALES**

|  | **Model 1** | | **Model 2** | |
| --- | --- | --- | --- | --- |
|  | **HR** | **95% CI** | **HR** | **95% CI** |
| **Anxiety/depression** |  |  |  |  |
| **ASD** | **2.39** | **2.10, 2.71** | **2.39** | **2.11, 2.72** |
| Deprivation** |  |  | 1.08 | 1.03, 1.13 |
| **Self-harm** |  |  |  |  |
| **ASD** | **2.65** | **2.12, 3.32** | **2.66** | **2.12, 3.33** |
| Deprivation** |  |  | 1.14 | 1.06, 1.24 |
| **Alcohol use** |  |  |  |  |
| **ASD** | **1.08** | **0.80, 1.46** | **1.08** | **0.80, 1.47** |
| Deprivation** |  |  | 1.13 | 1.03, 1.24 |
| **Drug use** |  |  |  |  |
| **ASD** | **1.98** | **1.46, 2.70** | **1.99** | **1.46, 2.70** |
| Deprivation** |  |  | 1.17 | 1.05, 1.31 |
| Emergency Department **use** |  |  |  |  |
| **ASD** | **0.90** | **0.84, 0.95** | **0.90** | **0.85, 0.95** |
| Deprivation** |  |  | 1.06 | 1.04, 1.08 |
| **Any primary care use** |  |  |  |  |
| **ASD** | **2.21** | **1.95, 2.49** | **2.21** | **1.96, 2.49** |
| Deprivation** |  |  | 1.08 | 1.03, 1.12 |
| **Any hospital use** |  |  |  |  |
| **ASD** | **0.91** | **0.90, 0.96** | **0.91** | **0.86, 0.97** |
| Deprivation** |  |  | 1.06 | 1.04, 1.08 |

Models incrementally adjusting for covariates. * Age at end of follow up period as time variable; ** WIMD quintile

Table s13: Cox’s regression analysis: **ASD associations with outcomes FEMALES**

|  | **Model 1** | | **Model 2** | |
| --- | --- | --- | --- | --- |
|  | **HR** | **95% CI** | **HR** | **95% CI** |
| **Anxiety/depression** |  |  |  |  |
| **ASD** | **1.78** | **1.50, 2.10** | **1.78** | **1.51, 2.10** |
| Deprivation** |  |  | 1.04 | 0.98, 1.10 |
| **Self-harm** |  |  |  |  |
| **ASD** | **3.60** | **2.68, 4.83** | **3.60** | **2.68, 4.83** |
| Deprivation** |  |  | 1.03 | 0.93, 1.15 |
| **Alcohol use** |  |  |  |  |
| **ASD** | **1.72** | **0.98, 3.02** | **1.72** | **0.98, 3.02** |
| Deprivation** |  |  | 1.01 | 0.84, 1.23 |
| **Drug use** |  |  |  |  |
| **ASD** | **4.99** | **2.07, 12.05** | **5.05** | **2.09, 12.19** |
| Deprivation** |  |  | 1.30 | 0.93, 1.81 |
| Emergency Department **use** |  |  |  |  |
| **ASD** | **1.16** | **1.04, 1.29** | **1.16** | **1.04, 1.29** |
| Deprivation** |  |  | 1.04 | 1.01, 1.08 |
| **Any primary care use** |  |  |  |  |
| **ASD** | **1.70** | **1.44, 2.01** | **1.70** | **1.44, 2.01** |
| Deprivation** |  |  | 1.03 | 0.98, 1.09 |
| **Any hospital use (inc ED)** |  |  |  |  |
| **ASD** | **1.15** | **1.04, 1.29** | **1.16** | **1.04, 1.29** |
| Deprivation** |  |  | 1.05 | 1.01, 1.08 |

Models incrementally adjusting for covariates. * Age at end of follow up period as time variable; **Proportion of follow up period with valid records ***WIMD quintile

Table s14: Cox’s regression – associations between ASD and outcomes, stratified by Welsh index of multiple deprivation (WIMD):

|  | **Model 1*** |  | **Model 2**** |  |
| --- | --- | --- | --- | --- |
|  | **HR** | **95% CI** | **HR** | **95% CI** |
| **Anxiety/depression** |  |  |  |  |
| WIMD 1: | 2.13 | 1.66, 2.72 | 2.17 | 1.70, 2.78 |
| WIMD 2: | 2.14 | 1.64, 2.79 | 2.22 | 1.70, 2.89 |
| WIMD 3: | 2.56 | 2.04, 3.22 | 2.45 | 1.95, 3.07 |
| WIMD 4: | 1.96 | 1.58, 2.42 | 2.02 | 1.63, 2.50 |
| WIMD 5: | 1.98 | 1.63, 2.40 | 2.00 | 1.65, 2.42 |
| **Self-harm** |  |  |  |  |
| WIMD 1: | 3.09 | 1.88, 5.09 | 3.17 | 1.93, 5.22 |
| WIMD 2: | 2.80 | 1.80, 4.37 | 2.91 | 1.86, 4.53 |
| WIMD 3: | 4.02 | 2.68, 6.04 | 3.81 | 2.54, 5.73 |
| WIMD 4: | 2.14 | 1.50, 3.06 | 2.20 | 1.53, 3.15 |
| WIMD 5: | 3.10 | 2.19, 4.38 | 3.12 | 2.20, 4.41 |
| **Alcohol use** |  |  |  |  |
| WIMD 1: | 1.45 | 0.73, 2.89 | 1.45 | 0.73, 2.88 |
| WIMD 2: | 1.04 | 0.53, 2.00 | 1.03 | 0.54, 1.99 |
| WIMD 3: | 1.48 | 0.80, 2.74 | 1.46 | 0.79, 2.70 |
| WIMD 4: | 0.74 | 0.40, 1.37 | 0.73 | 0.39, 1.35 |
| WIMD 5: | 1.45 | 0.89, 2.34 | 1.44 | 0.89, 2.32 |
| **Drug use:** |  |  |  |  |
| WIMD 1: | 1.53 | 0.65, 3.57 | 1.51 | 0.64, 3.53 |
| WIMD 2: | 2.12 | 1.01, 4.44 | 2.07 | 0.99, 4.34 |
| WIMD 3: | 3.34 | 1.52, 7.33 | 3.42 | 1.56, 7.52 |
| WIMD 4: | 2.45 | 1.45, 4.14 | 2.38 | 1.41, 4.02 |
| WIMD 5: | 1.88 | 1.09, 3.24 | 1.86 | 1.08, 3.22 |
| **ED Use:** |  |  |  |  |
| WIMD 1: | 0.94 | 0.82, 1.07 | 0.94 | 0.82, 1.07 |
| WIMD 2: | 0.96 | 0.83, 1.09 | 0.96 | 0.84, 1.09 |
| WIMD 3: | 1.03 | 0.92, 1.15 | 1.02 | 0.91, 1.15 |
| WIMD 4: | 0.90 | 0.80, 0.99 | 0.89 | 0.80, 1.00 |
| WIMD 5: | 0.96 | 0.86, 1.06 | 0.95 | 0.86, 1.06 |
| **Any primary care use:** |  |  |  |  |
| WIMD 1: | 1.96 | 1.54, 2.50 | 2.00 | 1.57, 2.54 |
| WIMD 2: | 1.88 | 1.47, 2.42 | 1.95 | 1.52, 2.51 |
| WIMD 3: | 2.38 | 1.91, 2.96 | 2.28 | 1.83, 2.85 |
| WIMD 4: | 1.94 | 1.58, 2.39 | 1.99 | 1.62, 2.45 |
| WIMD 5: | 1.89 | 1.57, 2.29 | 1.91 | 1.58, 2.30 |
| **Any hospital use (inc ED)** |  |  |  |  |
| WIMD 1: | 0.94 | 0.83, 1.07 | 0.94 | 0.83, 1.08 |
| WIMD 2: | 0.97 | 0.85, 1.10 | 0.96 | 0.85, 1.10 |
| WIMD 3: | 1.04 | 0.93, 1.17 | 1.03 | 0.92, 1.16 |
| WIMD 4: | 0.91 | 0.82, 1.01 | 0.91 | 0.81, 1.01 |
| WIMD 5: | 0.96 | 0.86, 1.07 | 0.95 | 0.86, 1.07 |

* Unadjusted association with ADHD; ** Association with ADHD, controlling for Sex and proportion of follow up.

Table s15: Cox’s regression analysis: **ASD associations with outcomes**

|  | **Model 1** | | **Model 2** | | **Model 3** | |
| --- | --- | --- | --- | --- | --- | --- |
|  | **HR** | **95% CI** | **HR** | **95% CI** | **HR** | **95% CI** |
| **Anxiety/depression** |  |  |  |  |  |  |
| **ASD** | **2.00** | **1.81, 2.22** | **2.02** | **1.82, 2.24** | **2.02** | **1.82, 2.24** |
| Sex |  |  | 0.46 | 0.41, 0.51 | 0.46 | 0.41, 0.51 |
| Deprivation** |  |  |  |  | 1.06 | 1.02, 1.09 |
| **Self-harm** |  |  |  |  |  |  |
| **ASD** | **2.96** | **2.45, 3.57** | **2.98** | **2.47, 3.60** | **2.98** | **2.47, 3.60** |
| Sex |  |  | 0.46 | 0.38, 0.56 | 0.47 | 0.38, 0.57 |
| Deprivation** |  |  |  |  | 1.08 | 1.01, 1.15 |
| **Alcohol use** |  |  |  |  |  |  |
| **ASD** | **1.27** | **0.96, 1.68** | **1.27** | **0.96, 1.68** | **1.27** | **0.96, 1.68** |
| Sex |  |  | 1.12 | 0.81, 1.54 | 1.13 | 0.82, 1.56 |
| Deprivation** |  |  |  |  | 1.10 | 1.00, 1.21 |
| **Drug use** |  |  |  |  |  |  |
| **ASD** | **1.95** | **1.43, 2.67** | **1.94** | **1.42, 2.65** | **1.94** | **1.43, 2.65** |
| Sex |  |  | 2.23 | 1.38, 3.60 | 2.27 | 1.41, 3.67 |
| Deprivation** |  |  |  |  | 1.19 | 1.07, 1.33 |
| Emergency Department **use** |  |  |  |  |  |  |
| **ASD** | **0.95** | **0.90, 1.00** | **0.95** | **0.90, 1.00** | **0.95** | **0.90, 1.00** |
| Sex |  |  | 0.97 | 0.92, 1.03 | 0.98 | 0.92, 1.03 |
| Deprivation** |  |  |  |  | 1.05 | 1.03, 1.06 |
| **Any primary care use** |  |  |  |  |  |  |
| **ASD** | **1.86** | **1.68, 2.06** | **1.87** | **1.69, 2.07** | **1.87** | **1.70, 2.07** |
| Sex |  |  | 0.49 | 0.44, 0.54 | 0.49 | 0.44, 0.54 |
| Deprivation** |  |  |  |  | 1.05 | 1.02, 1.09 |
| **Any hospital use (inc ED)** |  |  |  |  |  |  |
| **ASD** | **0.96** | **0.91, 1.01** | **0.96** | **0.91, 1.01** | **0.96** | **0.91, 1.01** |
| Sex |  |  | 0.97 | 0.92, 1.03 | 0.98 | 0.92, 1.03 |
| Deprivation** |  |  |  |  | 1.05 | 1.03, 1.06 |

Models incrementally adjusting for covariates. * Age at end of follow up period as time variable; **WIMD quintile

Supplementary table s16: Directly assessed ADHD hybrid cohort and e-cohort only sample comparisons:

|  | **Nested ADHD sample** | **e-cohort sample** | **OR (95% CI) or t** |
| --- | --- | --- | --- |
| **N** | 154 | 9379 |  |
|  | Mean (sd) or N (%) | |  |
| **Male gender** | 134 (87.0) | 7583 (80.90) | 1.587 (0.99, 2.55) |
| **Age at end of follow up (GP records)** | 19.11 (2.20) | 21.20 (3.80) | t=11.320, p<0.001 |
| **Deprivation index quintile** | 3.435 (1.56) | 3.421 (1.40) | t=-0.11, p=0.91 |
| **Age at ADHD diagnosis: Mean (sd)** | 8.41 (3.34) | 10.57 (3.86) | t= 7.95 p<0.001 |
| **Age at ASD diagnosis: Mean (sd)** | 9.33 (5.44) | 10.96 (5.09) | t=1.45, p=0.15 |
| **With comorbid ASD: N (%)** | 26 (16.90) | 1340 (14.30) | t=1.36, p=0.19 |
| **Anxiety/depression- any: N (%)** | 15 (9.70) | 1977 (21.10) | 0.40 (0.24, 0.69) |
| **Anxiety/depression events: Mean (sd)** | 0.14 (0.50) | 0.50 (1.45) | t=8.223, p=<0.001 |
| **Drug use - any: N (%)** | 10 (6.50) | 800 (8.50) | 0.745 (0.39, 1.42) |
| **Drug use - events: Mean (sd)** | 0.08 (0.34) | 0.27 (2.50) | t=0.15, p=0.36 |
| **Alcohol use – any: N (%)** | 5 (3.20) | 675 (7.20) | 0.43 (0.18, 1.06) |
| **Alcohol use – events: Mean (sd)** | 0.06 (0.37) | 0.19 (1.71) | t=0.93, p=0.35 |
| **Self-harm – any: N (%)** | 6 (3.9) | 1158 (12.3) | t=6.947, p<0.001 |
| **Self-harm – events: Mean (sd)** | 0.09 (0.50) | 0.40 (1.91) | 0.288 (0.13, 0.65) |
| **Emergency Department – any: N (%)** | 98 (63.6) | 6021 (64.2) | 0.976 (0.701, 1.36) |
| **Emergency Department** **– events: Mean (sd)** | 2.36 (3.81) | 3.53 (7.18) | 3.69, p<0.001 |

Supplementary figures:

Figure S1: Databases and process to identify directly assessed ADHD cohort:

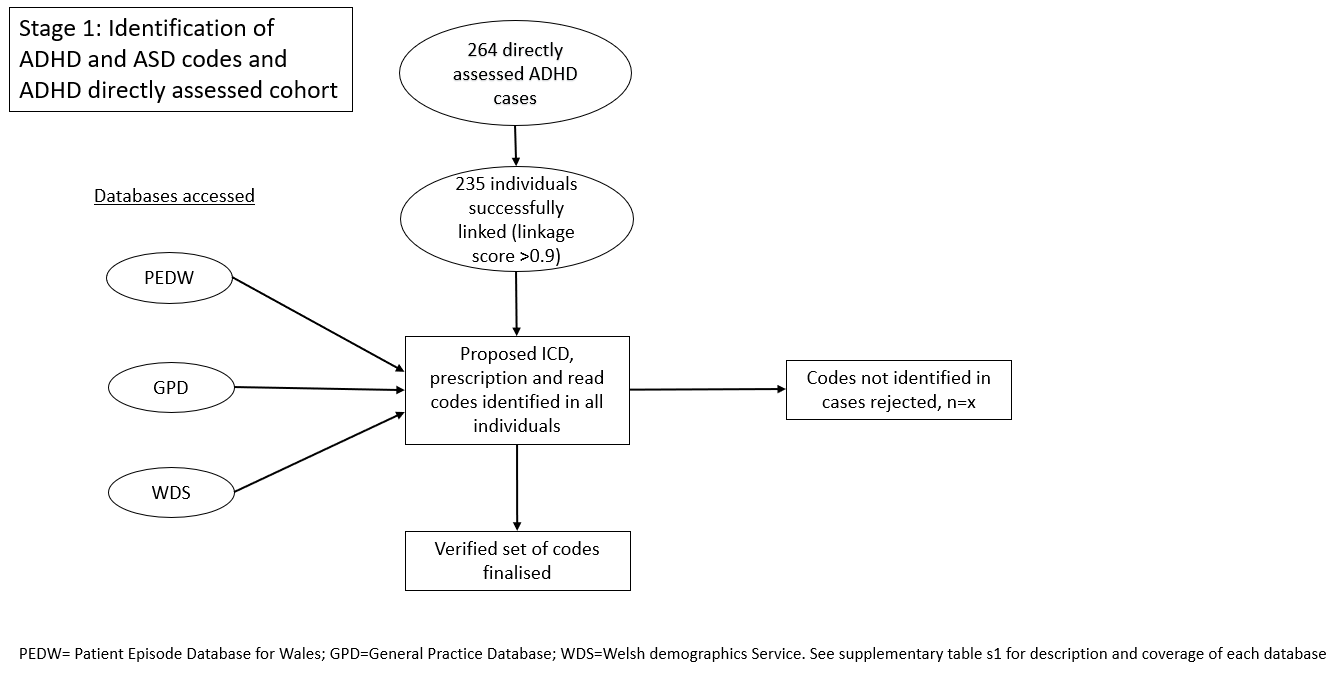

Figure S2: Databases and process to identify individuals with ADHD and ASD:

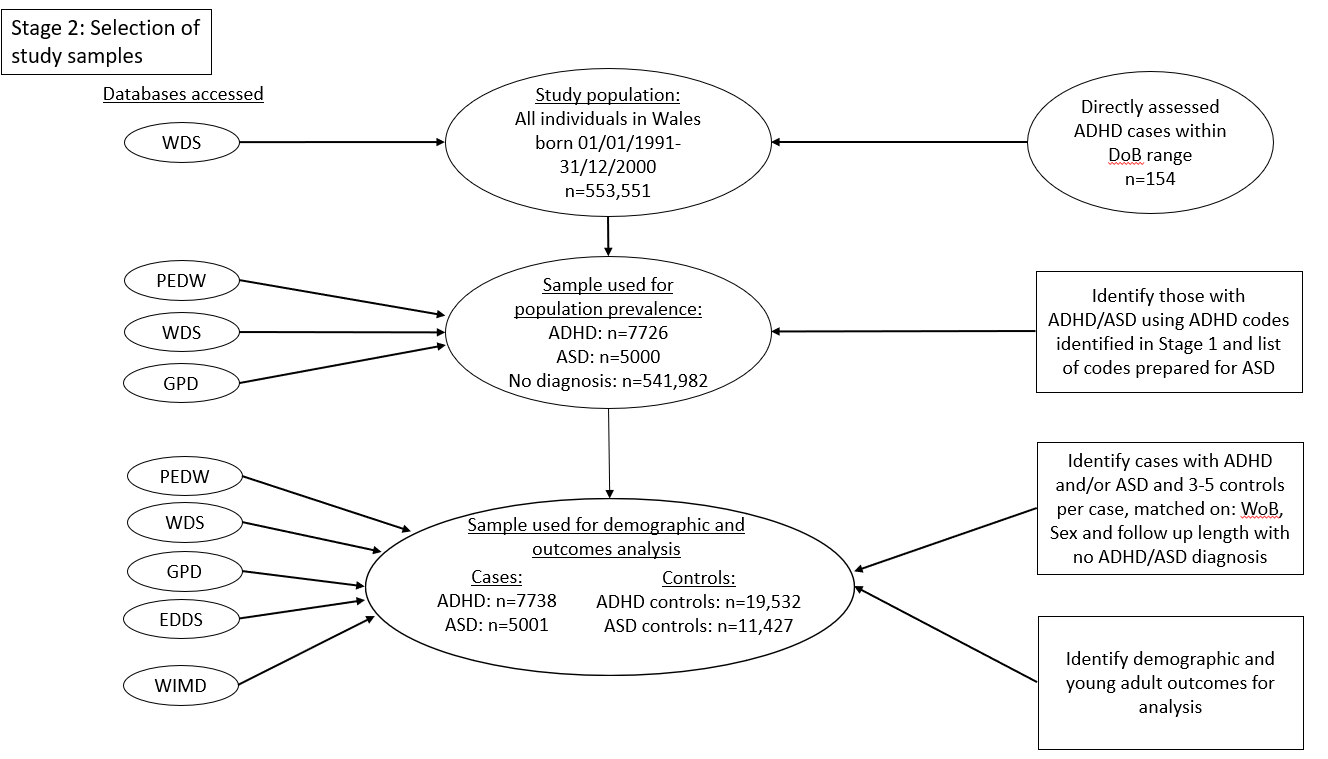

Figure S3: Associations between ADHD and early adult outcomes, by deprivation index:

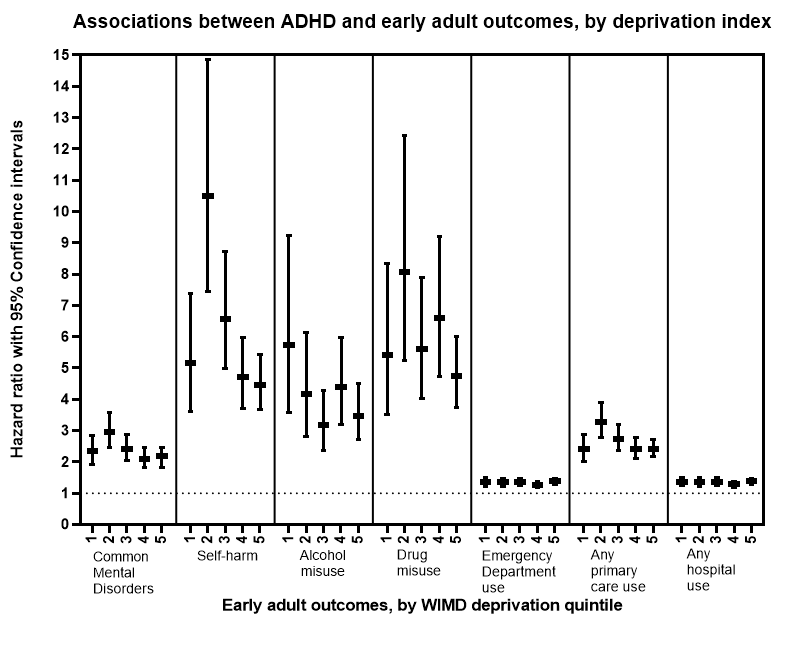

Supplementary Figure S4: Associations between ASD and early adult outcomes,:

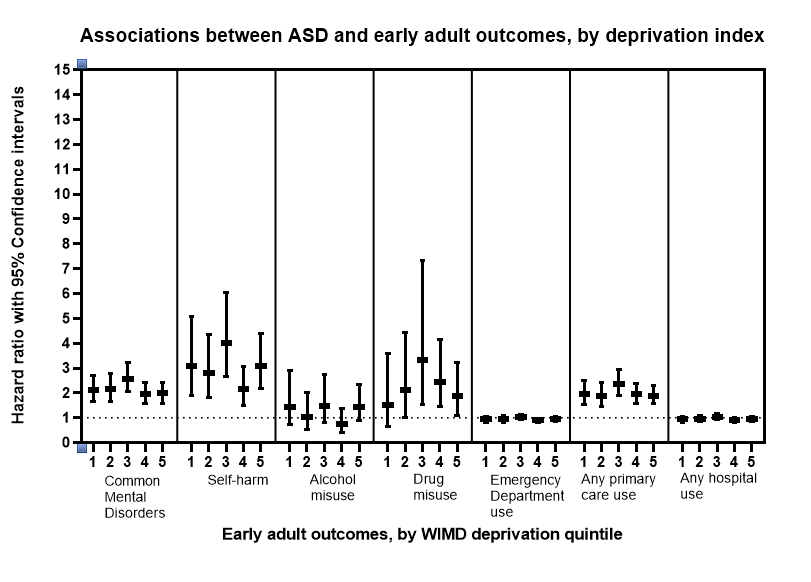

.
